# Tailored magnetic resonance fingerprinting of post-operative pediatric brain tumor patients

**DOI:** 10.1101/2022.09.22.22279737

**Authors:** Pavan Poojar, Enlin Qian, Zhezhen Jin, Maggie Fung, Alexis B Maddocks, Sairam Geethanath

**Affiliations:** Accessible Magnetic Resonance Laboratory, Biomedical Imaging and Engineering Institute, Department of Diagnostic, Molecular and Interventional Radiology, Icahn School of Medicine at Mount Sinai, New York, NY, United States; Columbia Magnetic Resonance Research Center, Columbia University, New York, NY, United States; Department of Biostatistics, Columbia University, New York, NY, United States; GE Healthcare Applied Sciences Laboratory East, New York, NY, United States; Columbia University Irving Medical Center, New York, NY, United States

## Abstract

**Purpose:** Brain and spinal cord tumors are the second most common cancer in children and account for one out of four cancers diagnosed. However, the long acquisition times associated with acquiring both data types prohibit using quantitative MR (qMR) in pediatric imaging protocols. This study aims to demonstrate the tailored magnetic resonance fingerprinting’s (TMRF) ability to simultaneously provide quantitative maps (T_1_, T_2_) and multi-contrast qualitative images (T_1_ weighted, T_1_ FLAIR, T_2_ weighted) rapidly in pediatric brain tumor patients.

**Methods:** In this work, we imaged five pediatric patients with brain tumors (resected/residual) using TMRF at 3T. We compared the TMRF-derived T_2_ weighted images with those from the vendor-supplied sequence (as the gold standard, GS) for healthy and pathological tissue signal intensities. The relaxometric maps from TMRF were subjected to a region of interest (ROI) analysis to differentiate between healthy and pathological tissues. We performed the Wilcoxon rank sum test to check for significant differences between the two tissue types.

**Results:** We found significant differences (P < 0.05) in both T_1_ and T_2_ ROI values between the two tissue types. A strong correlation was found between the TMRF-based T_2_ weighted and GS signal intensities for the healthy (correlation coefficient, r = 0.99) and pathological tissues (r = 0.88).

**Conclusion:** The TMRF implementation provides the two relaxometric maps and can potentially save ∼2 minutes if it replaces the T_2_-weighted imaging in the current protocol.

## INTRODUCTION

Over 4,000 brain and spinal cord tumors are reported yearly in children, and it is the second most common cancer and accounts for one out of four cancers diagnosed in children. Once diagnosed with brain tumors, three out of four children survive five years, depending on the tumor type, location, and related pathophysiology. [1]

Malignant tumors tend to proliferate and may regrow after being resected. MRI is ideal for neuro-oncology, especially for tumor localization and follow-ups. [2] However, MRI requires long acquisition times. Motion artifacts are most common in the pediatric population and lead to non-diagnostic brain MRI scans. To overcome this, children should be sedated during an MRI scan. [3] However, sedation is typically associated with long-term risks, adverse effects, and high costs. Rapid MRI has been employed to address different pediatric imaging needs. [4–7] These methods rely on accelerating data acquisition using parallel imaging [8], compressed sensing [9–13], non-Cartesian imaging [14], and simultaneous multi-slice imaging. [15]

### Quantitative imaging

Dixon et al. [16] analyzed 550 pediatric patients with brain tumors and showed the accuracy of qualitative diagnostic imaging. Consequently, quantitative MR (qMR) imaging plays a crucial role in diagnosing. [17–22] However, volumetric T_1_ and T_2_ maps are not considered in a routine clinical examination due to long scan times. [23] Recent methods have shown that relaxometry maps can be obtained rapidly. [23–28]

### Synthetic MR

Recently, synthetic contrasts have been generated using quantitative maps to reduce scan time. [27] However, compared to conventional fluid-attenuated inversion recovery (FLAIR) images, the synthetic FLAIR images have a low contrast-to-noise ratio. Fluid pulsation and phase encoding artifacts are observed in synthetic T_2_ weighted images. [27] Due to partial volume artifacts, the interface of cerebrospinal fluid (CSF) and brain parenchyma appears to be hyperintense in synthetic FLAIR images.

### Accelerated brain tumor relaxometry

T_1_ and T_2_ maps have long been employed to differentiate the tumor region from surrounding tissues. [29–33] De Blank P et al. [34] have demonstrated that the T_1_ and T_2_ quantitative maps obtained from magnetic resonance fingerprinting (MRF) help to differentiate tissue characteristics in pediatric brain tumors. Pirkl et al. [23] have demonstrated that their method can rapidly characterize disease in adult patients with brain tumors using T_1_ and T_2_ maps and synthetically generated multi-contrast contrast images. However, there are no pulse sequences that rapidly, simultaneously, and non-synthetically acquire qualitative and quantitative pediatric brain tumor imaging data. To that end, we leverage our tailored MRF (TMRF) sequence to demonstrate its utility in the context of scanning pediatric patients with pathological resected/residual brain tumors who underwent post-operative MR scanning. TMRF simultaneously rapidly provides quantitative maps (T_1_ and T_2_) and multi-contrast qualitative images (T_1_-weighted, T_1_ FLAIR, and T_2_-weighted) in one scan. [35–38] The quantitative maps help differentiate pathologically (resected/residual tumors) from healthy tissue. The routine pediatric brain tumor protocol includes T_2_-weighted images and can be potentially replaced by TMRF to reduce scan time. Also, the other two contrasts may provide additional information to the radiologist and do not require additional scan time.

## MATERIAL AND METHODS

### TMRF design

The repetition time (TR), flip angle (FA), and echo time (TE) trains were designed for 1000 time points as in the previous implementations. [38] All reconstructed images were visually inspected to pick three contrasts: T_1_-weighted, T1-FLAIR, and T_2_-weighted, in line with the simulations. Supplementary Figure 1 (a, b) shows the FA and TR schedule for the TMRF method, respectively. Supplementary Figure 1 (c) shows the EPG simulated magnetization evolutions for white matter (WM), gray matter (GM), CSF, and fat as in ref. [38]. The T_1_-weighted, T_1_ FLAIR and T_2_-weighted contrast correspond to the time-point 151 (green arrow), 127 (purple arrow), and 27 (yellow arrow), respectively. The ranges of T_1_ and T_2_, dictionary generation, simulation, acquisition, and reconstruction of TMRF are the same as in previous studies. [35,38–40]

### Patient population

We scanned five pediatric patients with brain tumors after obtaining consent from their parents or guardians as part of an institutional review board-approved study. We included male or female patients with a brain tumor (active, follow-up study, or follow-up after resection) between the ages of 8 and 18. Table 1 details the five pediatric patients’ tumor type, gender, tumor grade and location, slice number where the tumor was prominent, and brief treatment history.

**Table 1:** Patient details. Summary of five pediatric patients with brain tumors, including tumor type, age, gender, tumor grade, tumor location, slice number, and treatment history. All patients were between 8 and 18 years old and underwent routine MR scans. TMRF was included as an add-on sequence after obtaining consent from the patient’s parents/guardians. The slice number mentioned in the table corresponds to the slice where the tumor was prominently seen. These slices were selected after consulting the radiologist and used for ROI analysis. Out of five patients, one had a residual tumor, two had post-operative changes after the tumor was resected completely, and the tumor was removed entirely from the remaining two patients who did not have postoperative changes and underwent MR scan as a follow-up. The number of slices acquired for all the patients was 25 except for one patient (third), for which 20 slices were acquired; TMRF – tailored magnetic resonance fingerprinting, ROI – region of interest.

FIGURES
| Patient number | Tumor type | Gender | Tumor grade (WHO) | Tumor Location | Slice number | Description |
| --- | --- | --- | --- | --- | --- | --- |
| 1 | BRAFV600E (+) Glioblastoma | M | IV | left temporal lobe | 9 | Tumor has been resected through surgery, received radiation therapy and chemotherapy. There are post operative changes |
| 2 | Ependymoma | M | II | Intra ventricular | 8 | Tumor has been resected and currently under observation. Underwent targeted therapy |
| 3 | Pilocytic astrocytoma | M | I | Cerebellar | 17 | Tumor has been resected completely |
| 4 | SHH - Medulloblastoma | M | IV | Cerebellar | 9 | Tumor has been resected completely, received radiation therapy and chemotherapy. There are post operative changes |
| 5 | Pilocytic astrocytoma | M | I | Hypothalamic | 9 | Presence of residual tumor |

### MR scanning

All pediatric patients were scanned on a 3T GE Discovery MR750W with an eight-channel head coil. We included TMRF as an add-on sequence to the existing routine brain tumor protocol (Supplementary Figure 2). The second quadrant in Supplementary Figure 2 lists pre-contrast sagittal T_1_ volume, axial diffusion-weighted imaging (DWI), and axial SWAN sequences, while the third quadrant shows the post-contrast acquisitions. The first and fourth quadrants show the qualitative and quantitative imaging parts of the TMRF sequence, respectively. An additional gold standard (GS) T_2_-weighted sequence (vendor supplied) was scanned immediately after TMRF with the same resolution and number of slices. The TMRF sequence and GS were acquired pre-contrast. The acquisition parameters for TMRF: minimum TR/TE were 14.7/1.9 ms with a matrix size of 225×225, a field of view (FOV) of 225×225 mm^2^, and slice thickness of 5 mm. The average scan times for the reference T_2_-weighted sequence was ∼2 minutes, and the routine brain scan protocol was ∼35 minutes (pre- and post-contrast injection). The TMRF scan required 16 seconds per slice.

### Reconstruction

All qualitative images were reconstructed using MATLAB (The Mathworks Inc, MA). The sliding window method with a window size of 89 was used to reconstruct all 1000 images. All time-points were reconstructed for the first patient data to verify the time-points for all the three contrasts, with the previously selected time-points being selected based on the adult brain. After the time-points were identified for all the three contracts, reconstruction was performed only on these three time-points for the remaining four patients to accelerate the reconstruction process. The obtained images had a lower signal-to-noise ratio (SNR) compared to the adult data [38], especially T_2_-weighted images, due to the smaller volume of the pediatric brain (compared to adults) and under-sampled k-space data. The T_2_-weighted contrast occurs at the 27^th^ time-point. Eighteen images were zero-filled as the sliding window was designed from −45 to +44 images (total of 89), leading to an under-sampled k-space data.

For reconstructing quantitative maps, we used a DL approach based on DRONE [38,41] for reconstruction. We modified the sliding window method detailed in Cao et al. [42] by applying the sliding window to the magnetization evolution before the modified DRONE inference instead of computing the sliding window images. Further reconstruction details to obtain qualitative images and quantitative maps are presented in ref. [38].

### Image denoising and analysis

All reconstructed images from TMRF obtained for the first patient data (225×225 x25 slices x3 contrasts) were independently passed through four image denoising methods which are in-built functions in MATLAB. These filters were (i) MSF: median filter with a window size of 3×3 followed by image sharpening with a window size of 2×2, (ii) DLF: image denoising using a Deep Learning Toolbox from MATLAB [43], (iii) WNF: Wiener filter, (iv) WLF: denoising using wavelets. [44] All filtered images were visually compared (image contrast and image blurring) with the unfiltered TMRF images for T_1_-weighted and T_1_ FLAIR images, as GS images were not acquired. The unfiltered and filtered images were also quantitatively assessed using a no-reference image quality metric which included naturalness image quality evaluator (NIQE) and blind/referenceless image spatial quality evaluator (BRISQUE) for all the three contrast images on the 14^th^ slice (GM, WM, and CSF were seen). These metrics were calculated using inbuilt MATLAB functions to pick the optimal filter. The selected filter was applied to the remaining four patients’ data. The mean intensity values of T_2_-weighted images obtained from TMRF were compared with gold standard T_2_-weighted images for the slice(s) containing the tumor.

The quantitative maps were obtained using the modified DRONE with the filtered signal intensity images as input. The region of interest (ROI) to identify the tumor was drawn on the GS reference T_2_-weighted images with the help of the radiologist. First, the slice was selected where the pathology was clearly visible and then the ROI was drawn on that region. Another ROI considered healthy tissue was also drawn on the same slice. This procedure was repeated for all the patients’ GS T_2_-weighted images. The scanner output of the GS T_2_ was interpolated to 512×512.

Consequently, the ROI was drawn on the pathological and the healthy tissue on the T_1_ map obtained from TMRF on the corresponding slice. We also used these ROIs for the T_2_ map and T_2_-weighted data from the TMRF sequence. The mean and standard deviation (SD) of T_1_ and T_2_ values of the resected/residual tumor and the healthy tissue were calculated for all the five patients’ data. In the first patient’s case, ROI was drawn on a white matter to observe postoperative changes rather than from the resected tumor areas. Similarly, the normalized mean signal intensity and standard deviation of T_2_-weighted values of the resected/residual and healthy tissue ROI were calculated for GS and TMRF. Since the intensity ranges were different for GS and TMRF, all the weighted images were normalized before calculating the mean.

### Statistical analysis

The null hypothesis was that there was no significant difference between the pathological and the healthy tissues with respect to the dependent variable mean in this pediatric tumor population. A p-value smaller than 0.05 was considered statistically significant. We performed the Wilcoxon rank-sum test to compare the ROIs using GraphPad (GraphPad Software Inc, CA), similar to the analysis performed in ref. [34]. The two-tailed test with a confidence interval of 95% was used to compute the exact p-value. We performed this test to evaluate any difference between two independent groups (pathological and healthy tissues). We computed the mean ± SD of the pathological and healthy tissue ROIs. We computed these means for the T_1_ and T_2_ maps and the GS T_2_-weighted and TMRF T_2_-weighted images. We performed a linear regression analysis between the GS T_2_-weighted and TMRF T_2_-weighted images for pathological and healthy ROIs’ mean signal intensity values. The R-squared statistic value or the coefficient of determination was computed to determine the correlation between the two methods. Also, we computed the range of differences between pathological and healthy tissue for T_1_ and T_2_ values.

## RESULTS

### MR scanning

The TMRF sequence will potentially save ∼2 minutes if relaxometric maps were added to the existing protocol by replacing the existing T_2_-weighted scan. Given that the TMRF scan provides T_1_, T_2_ maps, and T_2_-weighted images, the additional time added to the existing protocol to acquire the maps if the GS T_2_-weighted were to be replaced with TMRF is ∼5 minutes, i.e., additional protocol time = acquisition time (Tacq) for TMRF – Tacq for T_2_ GS. This acquisition time is two minutes fewer than a conventional MRF acquisition with similar spatio-temporal resolutions and the additional benefit of the three non-synthetic contrasts.

### Reconstruction and image denoising

Supplementary Figure 3 shows the qualitative reconstructed images (T_1_-weighted, T_1_ FLAIR, and T_2_-weighted images) obtained before and after image denoising the first patient’s data. We observed that MSF possesses residual noise. However, denoising using the WNF and WLF shows better denoising and less blurring than the MSF. However, these images appeared blurred compared to DLF (shown with a yellow arrow in Supplementary Figure 3). Hence, DLF was chosen for all three contrasts after visual inspection. Table 2 (a-c) shows the results obtained from the two referenceless quantitative metrics, NIQE and BRISQUE, for T_1_-weighted, T_1_ FLAIR, and T_2_-weighted images. NIQE and BRISQUE values were lesser for DL-based denoising (column colored with green) than for the other three filters validating the visual inspection shown in Supplementary Figure 3.

**Table 2:** Qualitative image quality metric. Two no-reference image quality metrics that are part of Matlab (NIQE and BRISQUE) were used on all four filtered and unfiltered images for all three contrasts (a) T_1_-weighted, (b) T_1_ FLAIR, and (c) T_2_-weighted images. This evaluation was performed on one patient data on the 14^th^ slice. The output of MSF, DLF, WNF, and WLF correspond to the median filter, followed by image sharpening, DL-based denoising, wiener filter, and denoising using wavelets, respectively. Filter with less NIQE and BRISQUE values (filter 2 – column filled in green) were considered, and the same filter was used for all the other patient data; NIQE - Naturalness Image Quality Evaluator, BRISQUE - Blind/Referenceless Image Spatial Quality Evaluator, FLAIR - Fluid attenuated inversion recovery, DL - deep learning

| a) T<sub>1</sub> weighted |  |  |  |  |  |
| --- | --- | --- | --- | --- | --- |
|  | unfiltered | MSF | DLF | WNF | WLF |
| NIQE | 10.3 | 6.5 | 5.6 | 5.8 | 7.1 |
| BRISQUE | 44.4 | 29.6 | 14.4 | 38.3 | 37.1 |
| b) T<sub>1</sub> FLAIR |  |  |  |  |  |
| NIQE | 12 | 6.2 | 5.6 | 6.2 | 6.8 |
| BRISQUE | 44.7 | 29.3 | 20.3 | 41.4 | 40.1 |
| c) T<sub>2</sub> weighted |  |  |  |  |  |
| NIQE | 17.7 | 5.8 | 3.5 | 7 | 7.8 |
| BRISQUE | 48.1 | 32 | 26.7 | 40.3 | 43.4 |

Figure 1 shows the qualitative images of all five representative pediatric patients with a brain tumor. The blue and green ROIs on the GS T_2_-weighted (first row) images indicate the resected or residual tumor region and healthy appearing tissue, respectively, except for Patient 1 (first column) and Patient 4 (fourth column), where the blue ROI indicates the post-operative changes. We saw that fat was suppressed in T_2_-weighted images obtained from TMRF (shown with an orange arrow). This contrast was due to the 180° magnetization preparation pulse. This image contrast is in agreement with the results obtained from the simulation (Supplementary Figure 1). All images shown here are after passing through DLF. Figure 2 shows the quantitative T_1_ and T_2_ maps obtained from TMRF for the five pediatric patients. We observed that the resected/residual tumor region shows higher T_1_ and T_2_ values than healthy tissue. For Patient 2 (Ependymoma), it was challenging to draw the ROI as the resected tumor was part of the intra-ventricular region. It can be observed that T_1_ and T_2_ values were high (in the range of CSF) and hyperintense in T_2_-weighted images (Figure 1).

**Figure 1:**
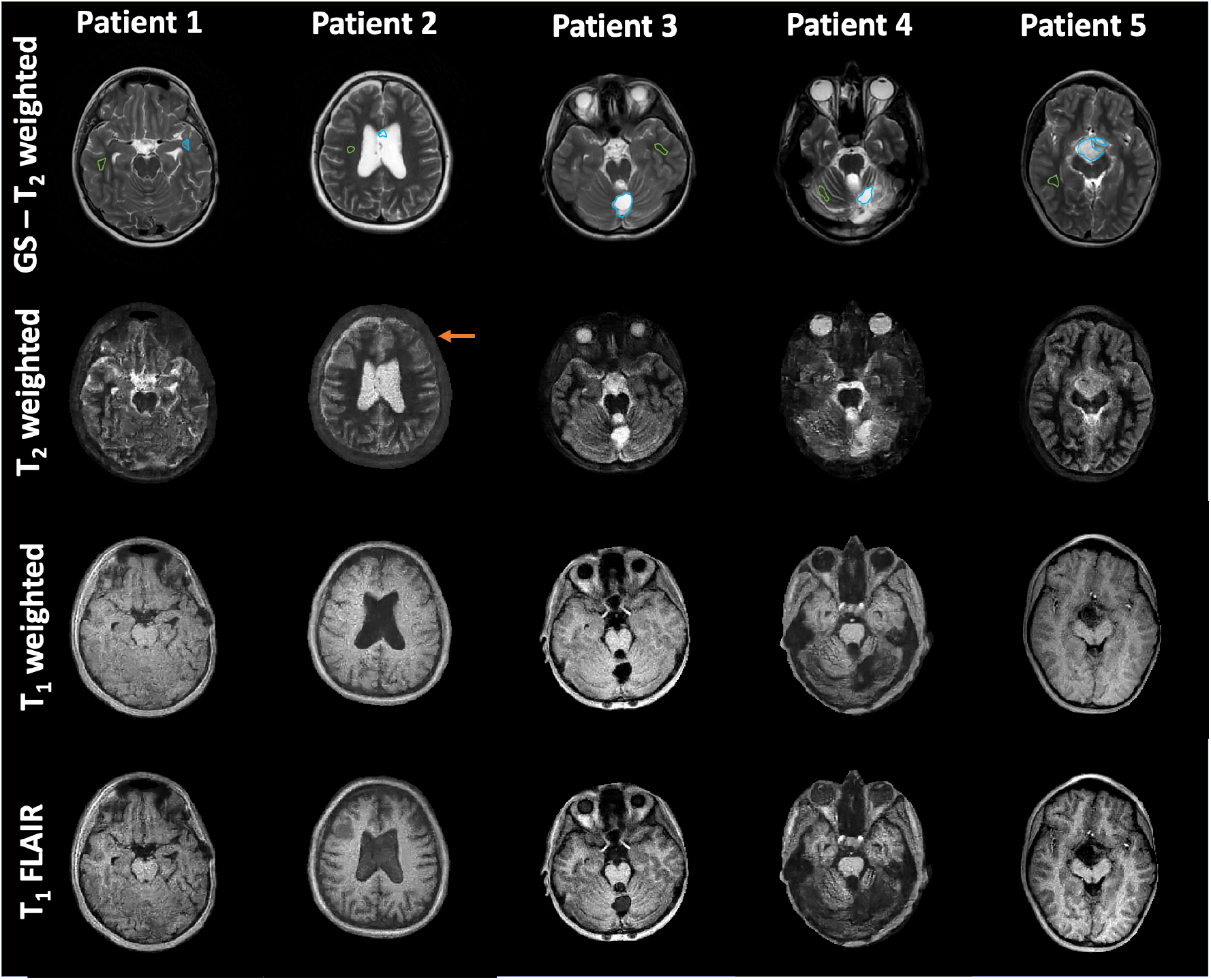
Qualitative study. Qualitative images of five representative pediatric patients were obtained from GS T_2_-weighted sequence (first row) and TMRF (second, third and fourth row). The tumors were resected for Patients 1, 2, and 3. However, Patient 1 and Patient 4 show some post-operative changes, as shown in blue ROI on GS T_2_-weighted image. Patient 5 has a residual tumor. The radiologist assisted in slice selection and drawing the first ROI on the GS images (blue ROI for resected/residual tumor and green ROI for healthy tissue). Henceforth, the same slice number and location were selected on the TMRF data, and ROI was drawn. All TMRF images shown here are DL denoised images. This procedure was followed for all the five patients’ data; GS – the gold standard, TMRF – tailored magnetic resonance fingerprinting, and ROI – a region of interest.

**Figure 2:**
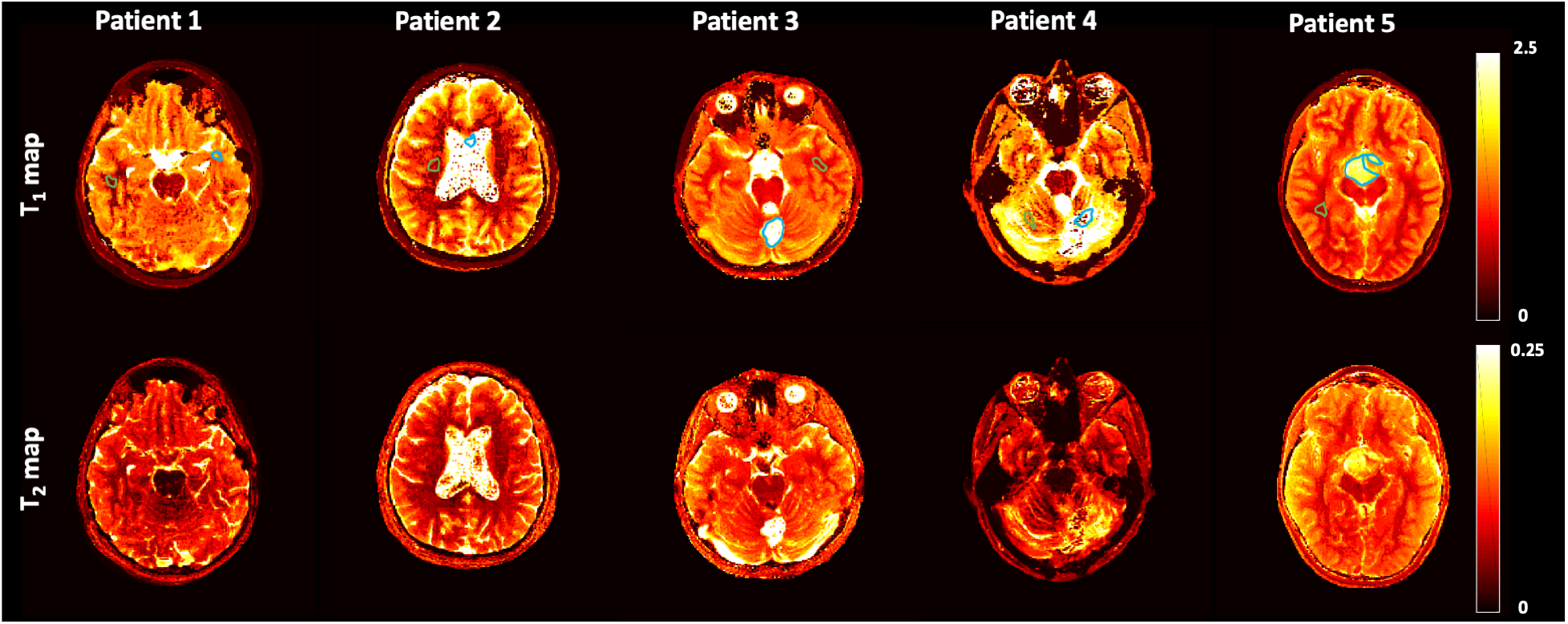
Quantitative study. T_1_ and T_2_ maps obtained from TMRF for five pediatric patients. More details about patients are included in Table 1. The ROI was drawn on the T_1_ map using reference ROI drawn on the GS T_2_-weighted images with the help of the radiologist. The same ROI (mask) was used on the T_2_ map and T_2_-weighted images (TMRF). The green ROI shows the healthy tissue drawn on the same slice. All maps shown here are after DL-based denoising, performed on the signal intensity (1000 images) before passing through the DRONE model. T_1_ and T_2_ values are in seconds; ROI - a region of interest, GS - gold standard, TMRF - tailored magnetic resonance fingerprinting, DRONE - MR fingerprinting deep reconstruction network.

### ROI and Statistical analysis

Figure 3 (a, b) illustrates the respective plot of mean T_1_ and T_2_ values of pathological and healthy tissue for all five patients (see Figure 2 with blue and green ROI). Figure 3 (c, d) shows the mean intensity values of pathological tumor and healthy tissue obtained from T_2_-weighted images of GS and TMRF, respectively (see Figure 1 with blue and green ROI). We observed from Figure 3 that the mean T_1_ and T_2_ values of pathological tumors are higher than healthy tissue for all five patients. These results are similar to the previously reported study. [34] Similarly, the mean intensity value for resected/residual tumors is higher than for healthy tissue (Figure 3 (c, d)). Also, the T_1_ and T_2_ differences between pathological and healthy tissue ranged from 500 ms to 2100 ms and 10 ms to 240 ms respectively.

**Figure 3:**
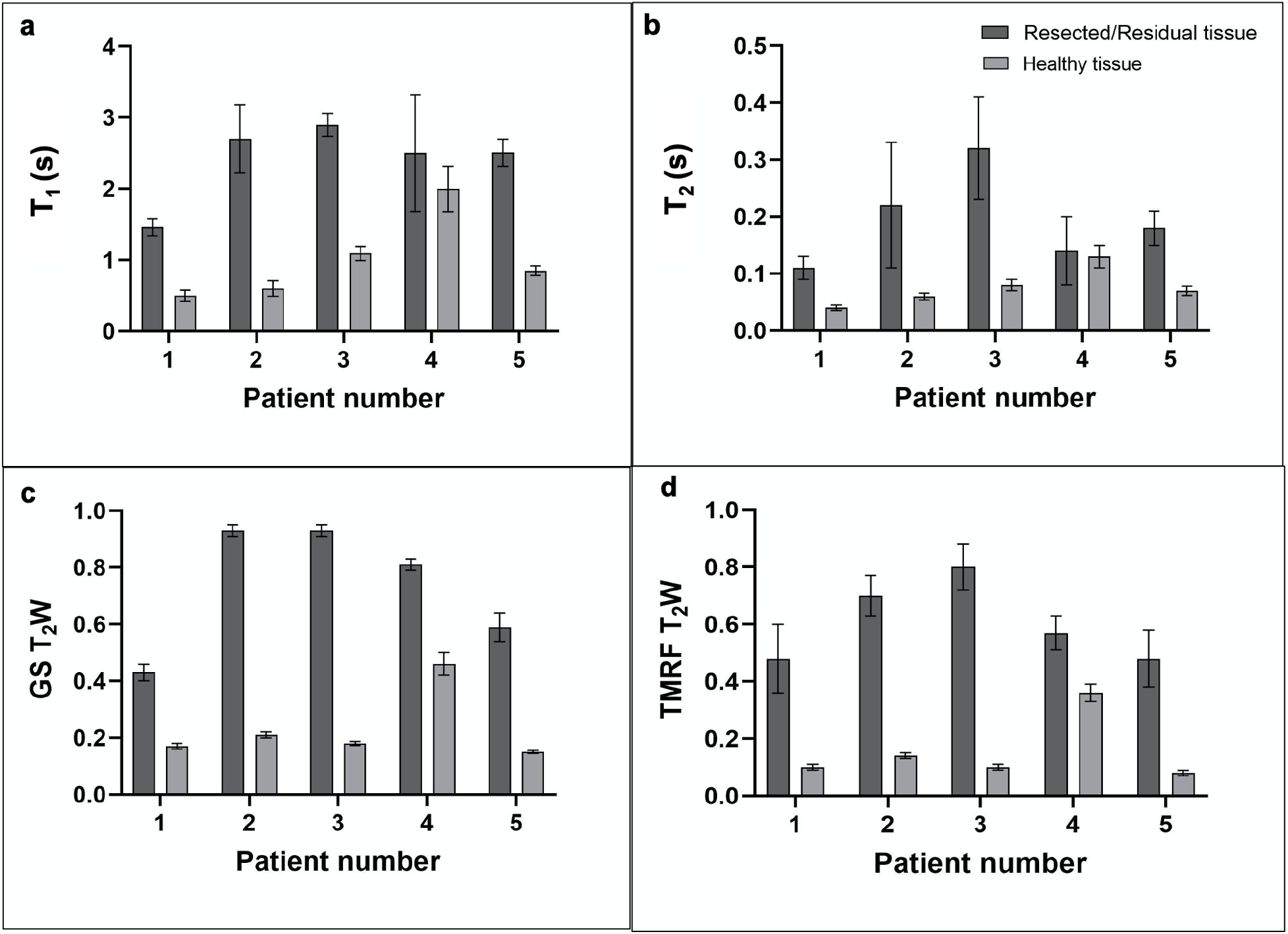
T_1_, T_2,_ intensity, and plots. The bar graph shows the mean and SD of (a) T_1_ and (b) T_2_ values of resected/residual tumor (dark gray) and healthy tissue (light gray) for all five patients. The second row shows the mean and SD of intensity values for (c) GS T_2_-weighted images and (d) TMRF T_2_-weighted images. The mean and SD were computed on the ROI, drawn manually on the GS T_2_-weighted (see Figure 1) and TMRF T_1_ map (see Figure 2). The intensity ranges of T_2_-weighted images from GS and TMRF were different. Hence, all qualitative images were normalized between 0 and 1. Out of the five patient data, two patients showed post-operative changes, and the ROI was drawn on these regions (see Table 1); SD - standard deviation, GS - the gold standard, TMRF - tailored magnetic resonance fingerprinting, ROI - a region of interest.

Supplementary Table 1 shows the detailed output of the Wilcoxon rank sum test performed using GraphPad. The p-values for the T_1_ map, T_2_ map, and GS T_2_-weighted data comparing the pathological and healthy ROIs were found to be 0.0159 for all three, and the p-value for TMRF T_2_-weighted was found to be 0.0079. As the p-values < 0.05, it was significant for both quantitative maps (T_1_ and T_2_) obtained from TMRF and qualitative T_2_-weighted images obtained from GS and TMRF. This shows that the pathological tissue region is significantly different from healthy tissue for the five patients’ data.

The correlation coefficients for healthy and pathological data from the GS T_2_-weighted and TMRF T_2_-weighted were 0.99 (R^2^ = 0.9984) and 0.88 (R^2^ = 0.7799). The linear regression plots are shown in Supplementary Figure 4 (a,b). These results indicate a strong correlation between the two methods with the relationship between the two methods for the healthy tissue being higher than the pathological tissue. However, these observations are limited by the small sample size of five patients.

## DISCUSSION AND CONCLUSION

This work focuses on post-operative pediatric brain tumor patients. An advantage of TMRF is that it takes about ∼16 seconds for one slice with better spatial resolution than the previous study (41 seconds per slice with a resolution of 1.17×1.17 mm^2^). [34] Also, TMRF has an additional advantage over non-synthetic contrast images. The resected/residual tumor can be seen in T_2_-weighted images from GS and TMRF (see Figure 1). As the T_2_-weighted sequence is part of routine pediatric brain tumor protocol, it can be potentially replaced by TMRF to reduce the total scan time. Since all contrast images and quantitative maps were obtained from a single scan, image registration challenges are expected to be minimal. Intra-scan motion is expected to distort the non-synthetic qualitative images, but the quantitative maps are less sensitive to motion. [26] The time-points for the three contrasts (T_1_ weighted, T_1_ FLAIR, and T_2_ weighted) are within the 200^th^ TR/FA combination (3.2 seconds per slice). This indicates that subject motion after this 200^th^ time-point will not affect the non-synthetic data as well on a slice-by-slice basis. This work can be extended to perform longitudinal studies on the patients where we can see the T_1_ and T_2_ changes in follow-up studies post-surgery or the effect of radiation therapy as discussed in ref. [19] or after chemotherapy. [45] Previous studies leverage the singular value decomposition (SVD) based method for TMRF image reconstruction for quantitative maps. [46,47] Previous studies show that MRF and TMRF are sensitive to differentiating T_1_ and T_2_ differences within physiological ranges. [38,47,48]

### Limitations

The slice thickness used in the study was 5 mm, which is based on previously published work by our group and others. [34] However, this needs to be reduced to 3 mm to meet the current clinical standards. The patient population has a small sample size (N = 5) with lower SNR images than GS. The TMRF implementation needs to be further optimized for pediatric scanning by allowing increased FA and TR values. Another limitation of TMRF is its reconstruction time. It takes ∼50 minutes for 25 slices over 1000 images as most of the reconstruction time was dedicated to Nonuniform fast Fourier transform (NUFFT) computation. For denoising, the current study used a pre-trained deep NN in MATLAB that assumes that the noise follows a Gaussian distribution with a limited range of SDs. Given that noise in MR images is hardware, pulse sequence-dependent, and typically follows a Rician distribution [49], we plan to develop a custom deep NN for denoising in the future that considers noise related to the TMRF acquisition similar to the approach in refs. [50] and [33]. Gold standard T_1_ and T_2_ maps also had a lesser SNR compared to the adult qMR data (similar to the qualitative data), which could be attributed to a smaller volume of the pediatric brain.

In conclusion, TMRF provides three contrasts (T_1_-weighted, T_1_ FLAIR, and T_2_-weighted) and two quantitative maps (T_1_ and T_2_) in one single scan. The T_1_ and T_2_ maps and pre-contrast T_2_-weighted images distinguish pathological (resected/residual tumor) from healthy tissue in post-operative pediatric brain tumor patients.

## Data Availability

All data produced in the present study are available upon reasonable request to the authors

## Acknowledgement

We would like to acknowledge Rolf F. Schulte for providing multi nuclear spectroscopy research pack (MNSRP); Guido Buonincontri and Pedro A Gomez for MRF package on MNSRP.

