## Supplementary materials for "Tailored magnetic resonance fingerprinting of post-operative pediatric brain tumor patients"

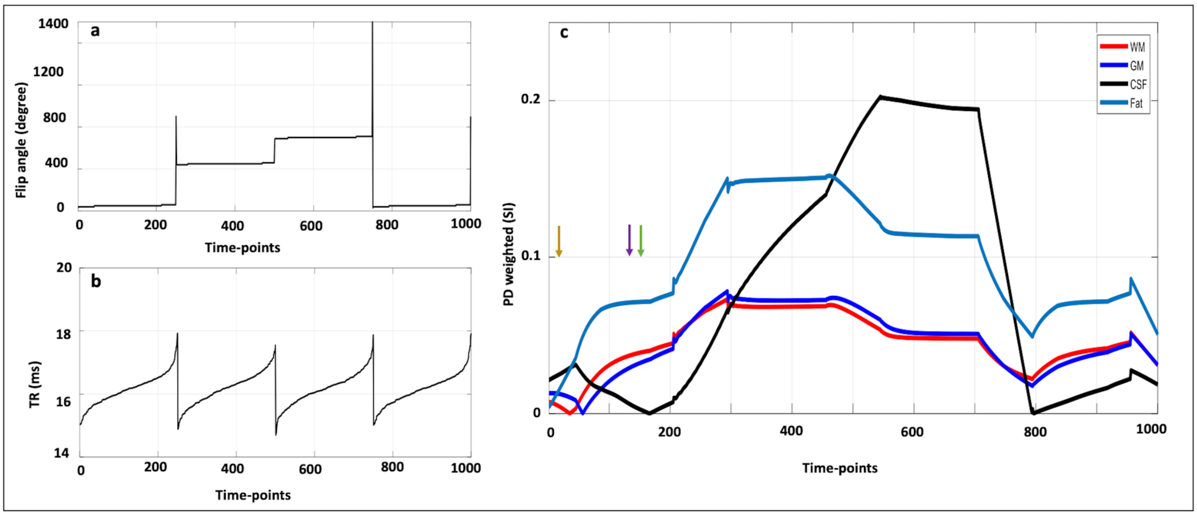


**Supplementary Figure 1:** ***TR, FA, and simulation*** – (a-c) TR, FA train, and EPG simulation for WM, GM, CSF, and fat for TMRF. The TR train consists of 1000 points with a minimum TR of 14.7 ms. The FA train includes 90° and 180° pulse at 250th and 750th time-points, respectively. TE was set to a minimum of 1.9 ms except between the 500th and 750th-time points. During these time-points, TE was modified to implement the 2-point Dixon method. Water and fat images were not used in this study). All 1000 images were reconstructed, and the three contrast images were selected for the first dataset. The time points were selected based on visual inspection and validated by the EPG simulated data. The yellow, purple, and green arrows show the selected time points for $T_2$-weighted, $T_1$-weighted, and $T_1$ FLAIR contrasts, respectively. From the second dataset onwards, the sliding window method was applied only on these three-time points to save reconstruction time. TR – repetition time, FA – flip angle, EPG – extended phase graph, WM – white matter, GM – gray matter, CSF – cerebrospinal fluid, TMRF – tailored magnetic resonance fingerprinting, FLAIR - Fluid attenuated inversion recovery.

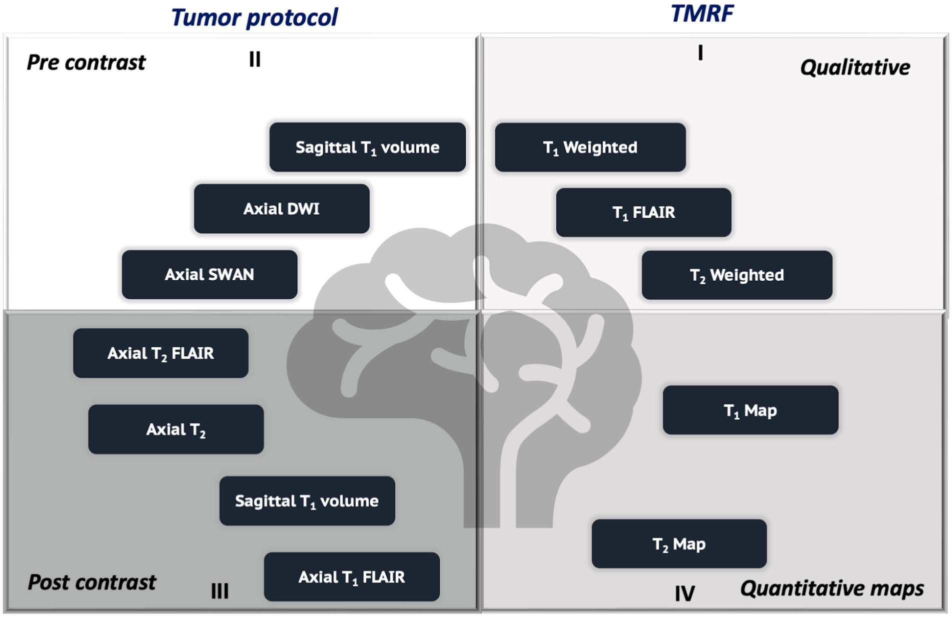


**Supplementary Figure 2:** ***Pediatric brain tumor protocol*** – Routine pediatric brain tumor protocol used in 3T GE Discovery MR750W clinical scanner. Pre-contrast sequences include sagittal $T_1$ volume, axial DWI, and axial SWAN (quadrant II). The post-contrast injection sequences include axial $T_2$ FLAIR, axial $T_2$-weighted, sagittal $T_1$ volume, and $T_1$ FLAIR (quadrant III). Images obtained from TMRF include $T_1$-weighted, $T_1$ FLAIR, and $T_2$-weighted contrasts (quadrant I) along with $T_1$ and $T_2$ maps (quadrant IV). TMRF was included as an add-on sequence to the routine brain protocol before the contrast injection after obtaining consent from the patient's parents/guardians. The total scan time for the routine pediatric brain tumor protocol was ~35 minutes (pre and post-contrast injection). TMRF takes 16 seconds per slice with a resolution of 1.1 x 1.1 x 5 $mm^3$. In addition to TMRF, resolution and slice number matched GS $T_2$-weighted (pre-contrast) was also acquired using the vendor-supplied sequence. This additional scan was to help localize the tumor; DWI - diffusion-weighted imaging, FLAIR - fluid-attenuated inversion recovery, TMRF – tailored magnetic resonance fingerprinting, GS – gold standard.

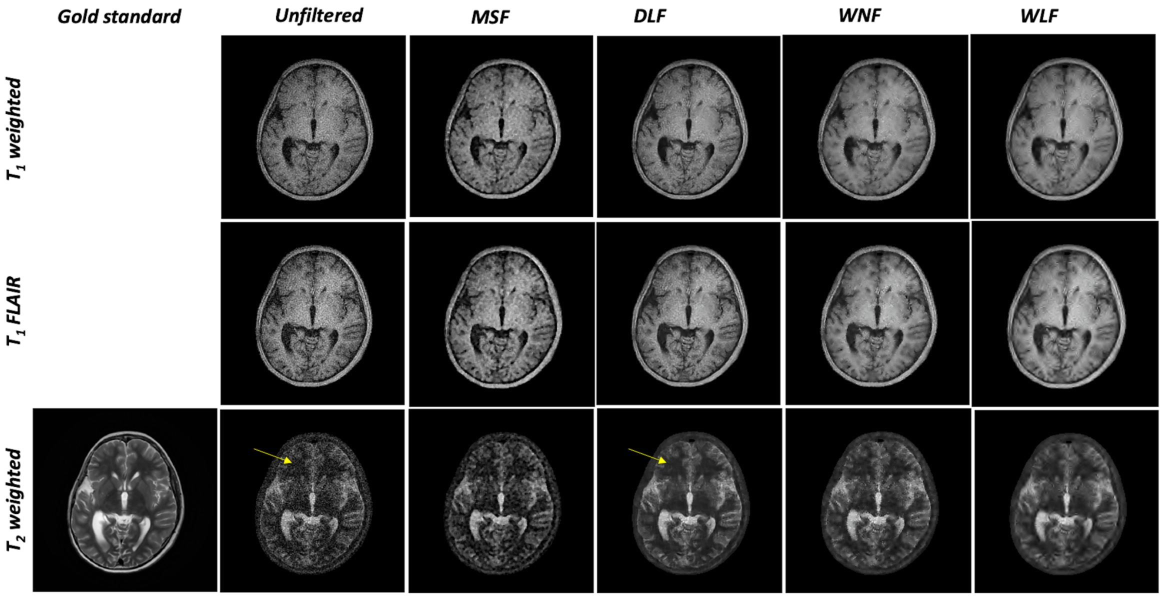


**Supplementary Figure 3:** ***Qualitative denoising*** –Four denoising filters that are in-built Matlab functions were used. The output of MSF corresponds to the median filter, followed by image sharpening. Similarly, DLF, WNF, and WLF outputs correspond to DL-based denoising, wiener filter, and denoising using wavelets. All four filtered outputs were visually compared with unfiltered images for $T_1$-weighted and $T_1$ FLAIR contrasts. $T_2$-weighted filtered images were compared with vendor-supplied GS $T_2$-weighted images (first column); TMRF -tailored magnetic resonance fingerprinting, FLAIR - Fluid attenuated inversion recovery, DL - deep learning, GS – gold standard.

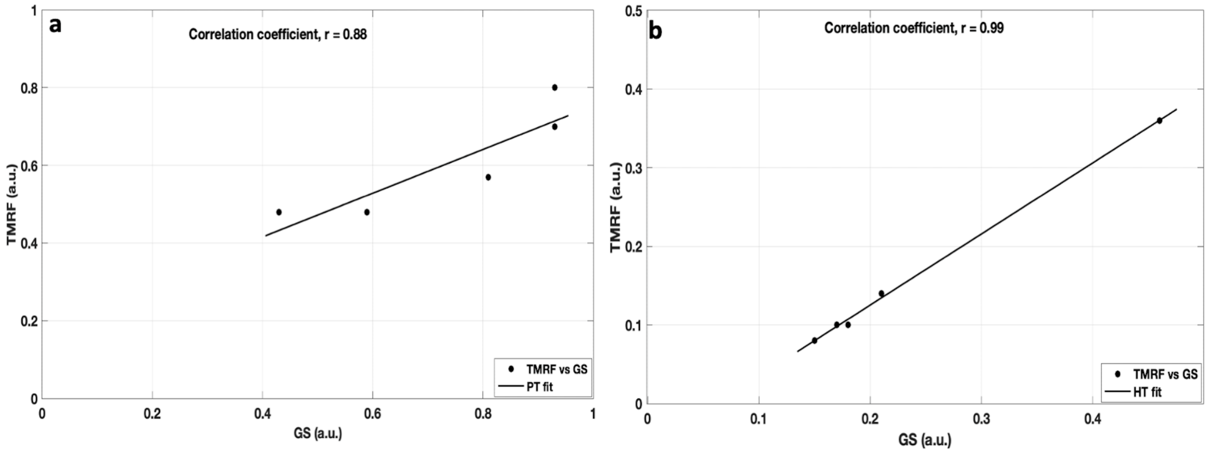


**Supplementary Figure 4:** ***Correlation between GS and TMRF $T_2$-weighted imaging*** – The linear regression fit of $T_2$-weighted images obtained from GS and TMRF for (a) PT and (b) HT. The ROI was drawn on the PT and HT on one slice where the PT was seen prominently. The correlation coefficient, r, for PT and HT was 0.88 and 0.99, respectively. GS - gold standard, TMRF – tailored magnetic resonance fingerprinting, PT - pathological tissue, HT - healthy tissue.

| $T_1$ map | | $T_2$ map | | GS $T_2$ weighted | | TMRF $T_2$ weighted | |
|---|---|---|---|---|---|---|---|
| P value | 0.0159 | P value | 0.0159 | P value | 0.0159 | P value | 0.0079 |
| Exact or approximate P value? | Exact | Exact or approximate P value? | Exact | Exact or approximate P value? | Exact | Exact or approximate P value? | Exact |
| P value summary | * | P value summary | * | P value summary | * | P value summary | ** |
| Significantly different (P < 0.05)? | Yes | Significantly different (P < 0.05)? | Yes | Significantly different (P < 0.05)? | Yes | Significantly different (P < 0.05)? | Yes |
| One- or two-tailed P value? | Two-tailed | One- or two-tailed P value? | Two-tailed | One- or two-tailed P value? | Two-tailed | One- or two-tailed P value? | Two-tailed |
| Sum of ranks in column A,B | 39 , 16 | Sum of ranks in column A,B | 39 , 16 | Sum of ranks in column A,B | 39 , 16 | Sum of ranks in column A,B | 40 , 15 |
| Mann-Whitney U | 1 | Mann-Whitney U | 1 | Mann-Whitney U | 1 | Mann-Whitney U | 0 |
| Difference between medians | | Difference between medians | | Difference between medians | | Difference between medians | |
| Median of column A | 2.510, n=5 | Median of column A | 0.1800, n=5 | Median of column A | 0.8100, n=5 | Median of column A | 0.5700, n=5 |
| Median of column B | 0.8500, n=5 | Median of column B | 0.07000, n=5 | Median of column B | 0.1800, n=5 | Median of column B | 0.1000, n=5 |
| Difference: Actual | -1.66 | Difference: Actual | -0.11 | Difference: Actual | -0.63 | Difference: Actual | -0.47 |
| Difference: Hodges-Lehmann | -1.65 | Difference: Hodges-Lehmann | -0.1 | Difference: Hodges-Lehmann | -0.47 | Difference: Hodges-Lehmann | -0.43 |

**Supplementary Table 1:** ***Statistical analysis*** – The Wilcoxon Rank Sum Test results on the $T_1$ map, $T_2$ map, GS $T_2$-weighted images and TMRF $T_2$-weighted images. The Wilcoxon rank sum test comparing the resected/residual tumor and healthy tissues: the $T_1$ map (p=0.016), $T_2$ map (p=0.016), GS $T_2$-weighted images (p=0.016) and TMRF $T_2$-weighted images (p=0.008). In all the cases, the p-value was <0.05, which shows that the resected/residual tumor is significantly different from healthy tissue; GS - the gold standard, TMRF - tailored magnetic resonance fingerprinting.
